# Capturing Trial-by-Trial Variability in Behaviour: People with Parkinson’s Disease Exhibit a Greater Rate of Short-Term Fluctuations in Response Times

**DOI:** 10.1101/2024.10.10.24315225

**Authors:** Hayley J. MacDonald, Ole Bernt Fasmer, Olav T. Jønsi, Lin Sørensen

## Abstract

Average response time is frequently used to reflect executive function. Less often studied is intra-individual variability in response times (IIVRT) which reflects within-person consistency. Higher IIVRT in Parkinson’s disease (PD) has been associated with poor executive function but almost exclusively studied using standard deviations. Such linear measures cannot capture rapid and spontaneous changes in biological systems such as dopaminergic bursting activity. Therefore, nonlinear measures may provide important complementary insights into dopamine-related neurocognition. Our primary aim was to investigate nonlinear IIVRT measures in PD using graph theory, constituting the first use of this approach on RT data. As hypothesized, PD was associated with a greater rate of trial-by-trial IIVRT compared to healthy older adults. These novel results indicate that a similarity graph algorithm may be a useful tool to capture the more rapidly varying and spontaneous changes in RT behavior that result from the dysfunctional dopamine bursting dynamics present in PD.

## Introduction

It is increasingly acknowledged that Parkinson’s disease (PD) results from a complicated interplay of genetic and environmental factors affecting numerous fundamental cellular processes (Kalia & Lang, 2015). PD is therefore increasingly recognized as more than purely a motor-deficit condition and includes varying degrees of cognitive impairment. Measures of response time (RT) are frequently used to investigate cognitive impairment. Average RT following presentation of an external stimulus reflects levels of attention, processing speed and executive function (MacDonald et al., 2006). Less often studied is the within-person consistency in RT. Higher intra-individual variability in response times (IIVRT) is sensitive to poor brain health (Haynes et al., 2017; MacDonald et al., 2006), predicts all-cause mortality in longitudinal studies (Batterham et al., 2014; Der & Deary, 2018), even when controlling for dementia and age-related cognitive decline (Kochan et al., 2017), and is a promising biomarker for neurogenerative diseases (Costa et al., 2019; Haynes et al., 2017; Roalf et al., 2016). Higher IIVRT is found in various conditions associated with executive dysfunction such as Alzheimer’s disease (Burton et al., 2006), schizophrenia (Pappa et al., 2021) and Attention-Deficit/Hyperactivity Disorder (Kofler et al., 2013). Several causal explanations for higher IIVRT in these conditions have been suggested, such as poor motor inhibition (Mostofsky & Simmonds, 2008), worse attentional alertness (Kuntsi & Stevenson, 2001), and deficiencies in temporal processing (Castellanos & Tannock, 2002; Toplak et al., 2006) - all dysfunctions present in PD. Indeed, studies have found greater IIVRT in PD compared to healthy controls (Burton et al., 2006; de Frias et al., 2007; Dujardin et al., 2013; Morrison et al., 2021).

IIVRT can be assessed through different measures. Higher IIVRT in PD has almost exclusively been shown using standard deviation (SD). SD provides a standardised measure of inconsistency in RTs but is necessarily calculated as an average measure across the entire RT distribution, precluding any trial-level investigation. Furthermore, linear methods may not be suitable to capture rapid and spontaneous changes in biological systems. Most importantly for PD, dopamine is discharged in bursts (Grace & Bunney, 1984), which may manifest as these types of transient changes in behaviour. Dopamine bursting activity is increasingly becoming the focus of other modalities e.g. electroencephalography (Shin et al., 2017). The bursting dynamics of dopamine are also relevant when measuring the effect of dopamine depletion on RT patterns. Therefore nonlinear measures may provide important complementary insights into neurocognition (Iconaru et al., 2022). In other dopamin-related disorders such as ADHD, calculating the ex-gaussian distribution of RTs through the nonlinear measure of tau has increased the understanding of RT fluctations by capturing the exponential component of the distribution (Leth-Steensen et al., 2000). Tau specifically indicates the mean of slower RTs. One study has compared IIVRT-tau in PD versus healthy controls but found no difference (Pappa et al., 2021). Tau warrants further investigation, drawing from literature on other dopamine-related conditions to consider what underlying mechanisms could be reflected by this measure in the context of PD.

The nonlinear method of graph theory is one viable approach to capture the complexity of the biological changes in PD and their effect on behaviour. There are other possible nonlinear measures such as those based on theories of chaos, fractality, and complexity (Fasmer et al., 2016). However, these measures are more challenging to interpret and therefore not widely used. The similarity graph algorithm has been used to complement average variability measures of inter-beat-intervals of the heart in ADHD (Kvadsheim et al., 2022), to understand connectivity dynamics in neuroimaging (Yu et al., 2018), and applied in studies of motor activity in schizophrenia and depression (Fasmer et al., 2018). The algorithm can assess several indices familiar from graph theory in relatively short time windows, reflecting moment-to-moment consistency across trials. Graph theory analysis therefore enables the measurement of RT consistency across smaller time windows, bringing us closer to elucidating the role of transient dopamine bursts in RT variability. Graph theory approaches have been used to study brain network connectivity in PD (Vecchio et al., 2021). However, to our knowledge, we are the first to apply this method to investigate RT data.

The aim of the current study was to increase the understanding of RT fluctuations in PD beyond the use of IIVRT-SDs. Nonlinear measures of RT patterns were studied by calculating tau and graph theoretical measures. IIVRT in PD has previously been investigated using externally cued tasks e.g. simple reaction time (Morrison et al., 2021) or Eriksen flanker tasks (Pappa et al., 2021). We employed a version of the anticipatory response inhibition task (ARIT) (He et al., 2022) which requires the anticipation and correct timing of an internally-generated response. This enabled us to additionally investigate the influence of predictive timing processes on IIVRT. People with PD find internally-generated responses more difficult than externally cued ones (Jahanshahi et al., 1995), likely reflecting that these internal timing processes are more sensitive to dysfunctional dopamine fluctuations and by extension RT variability. We hypothesised that the ARIT would reveal reduced similarity of RTs in PD versus healthy controls across the time windows investigated with the graph theory analysis. We further hypothesised that this would extend to higher IIVRT for people with PD in the average distribution measures of SD and tau.

## Methods

### Participants

Data used in the current study were collected as part of previous studies using the same behavioural task with 30 healthy older adults (MacDonald et al., 2016) (average age 60 years, 17 female) and 85 individuals with Parkinson’s disease (Hall et al., 2023) (average age 64 years, 36 female). This combined sample size was above the 98 participants required (G*Power 3.1) to detect a medium effect size between groups with 0.8 power for the primary dependent measures from graph theory analysis (post-hoc power values achieved for the two primary dependent measures were 0.98 and 0.99, see Results). Participants with Parkinson’s disease (PwPD) completed data collection while on their routine dopamine medication. Data were collected and ethical approval was obtained at the University of Auckland (New Zealand) and University of Birmingham (United Kingdom), and written informed consent was obtained from each participant. Inclusion criteria in both studies included being aged 40 – 80 years, no history of neurological illness (apart from Parkinson’s disease), and normal or corrected-to-normal vision. The study was not preregistered.

### Behavioural Data – Anticipatory Response Inhibition Task (ARIT)

All participants completed the computerized ARIT written in Matlab (The MathWorks, Natick, MA; MacDonald et al., 2016) or Inquisit 6 Lab (Version 6.5.1, Millisecond Software; Hall et al., 2023). At the start of each trial, participants were presented with a grey screen containing two while vertical bars and a horizontal black target line 4/5 towards the top of the bars (Figure 1). The trial began after a variable delay (400 – 900 ms) once participants held down both response keys using their left and right index fingers. Black indicators then started rising at equal rates within the two white bars; the left/right indicator controlled via the left/right index finger. The indicators reached the target line in 800 ms and terminated their rise in 1000 ms, unless halted prior by lifting either or both index fingers.

**Fig. 1.**
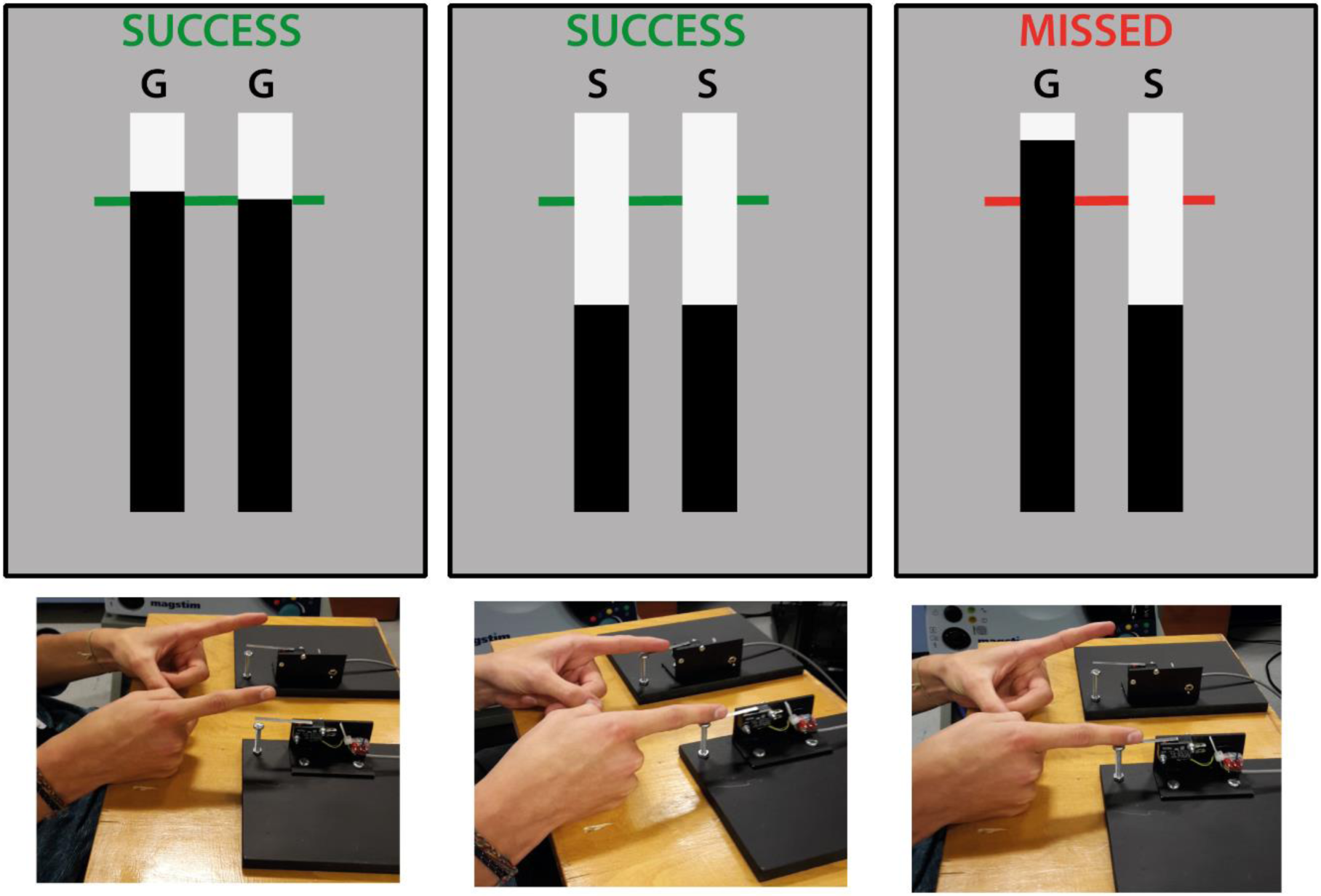
Experimental setup for behavioural task (MacDonald et al., 2021). The anticipatory response inhibition task display (top) and participant response (bottom) for a Go (left), Stop Bimanual (middle), and Stop Unimanual trial (right). The participant has successfully lifted from both switches (GG: Go, Go), kept both switches depressed (SS: Stop, Stop) and lifted from only the left-hand switch (GS: Go, Stop), respectively. Other type of Stop Unimanual trial (SG: Stop, Go) not shown.

The default response on Go trials required participants to intercept the rising indicators with the horizontal target line by correctly timing the lift of both fingers. On trial completion, visual feedback and a green target line indicated “success” (both releases within 30 ms of the target, Figure 1 left) or “miss” and a red target line to emphasize Go trials were to be performed as accurately as possible. The remaining trials were Stop trials, consisting of Stop Bimanual and Stop Unimanual trials. On Stop Bimanual trials participants had to cancel the bimanual lift response when both indicators stopped rising before reaching the target (Figure 1 middle). On Stop Unimanual trials either the left or right indicator stopped rising before reaching the target, requiring participants to cancel the lift response on the corresponding side, while still trying to intercept the remaining indicator at the target line (Figure 1 right). A staircase algorithm was used to converge on a 50% success rate for each Stop trial type by adjusting difficulty based on individual Stop trial performance (i.e. increasing/decreasing the indicator stop time by 25 ms on the next Stop trial if the previous trial was successful/unsuccessful).

Both task versions ensured a 2:1 Go:Stop trial ratio. Go trials were the primary trials of interest for the current study. Healthy controls (HC) performed 248 trials in total, consisting of 170 Go trials. PwPD completed 400 trials in total, consisting of 295 Go trials. For comparison between groups, the first 170 Go trials were analysed for PwPD.

### Dependent Measures

Response times (RTs) were calculated for Go trials (reported in milliseconds relative to trial onset) and averaged. To retain maximal variability data for each participant, RTs outside ± 3 standard deviations (SD) of their mean were replaced with the appropriate mean ± 3SD RT value (Kvadsheim et al., 2022) and any Go omissions (i.e. finger not lifted within 200 ms of the target) were replaced with the maximum RT of 1000 ms possible on Go trials (Verbruggen et al., 2019). To measure IIV across the entire Go RT distribution, SD (gaussian) and tau (ex-gaussian) were calculated for each participant (Figure 2A&B). Tau was calculated as [τ = SD * (skewness / 2) (1/3)] and reflects the positive skew of the RT distribution.

**Fig. 2.**
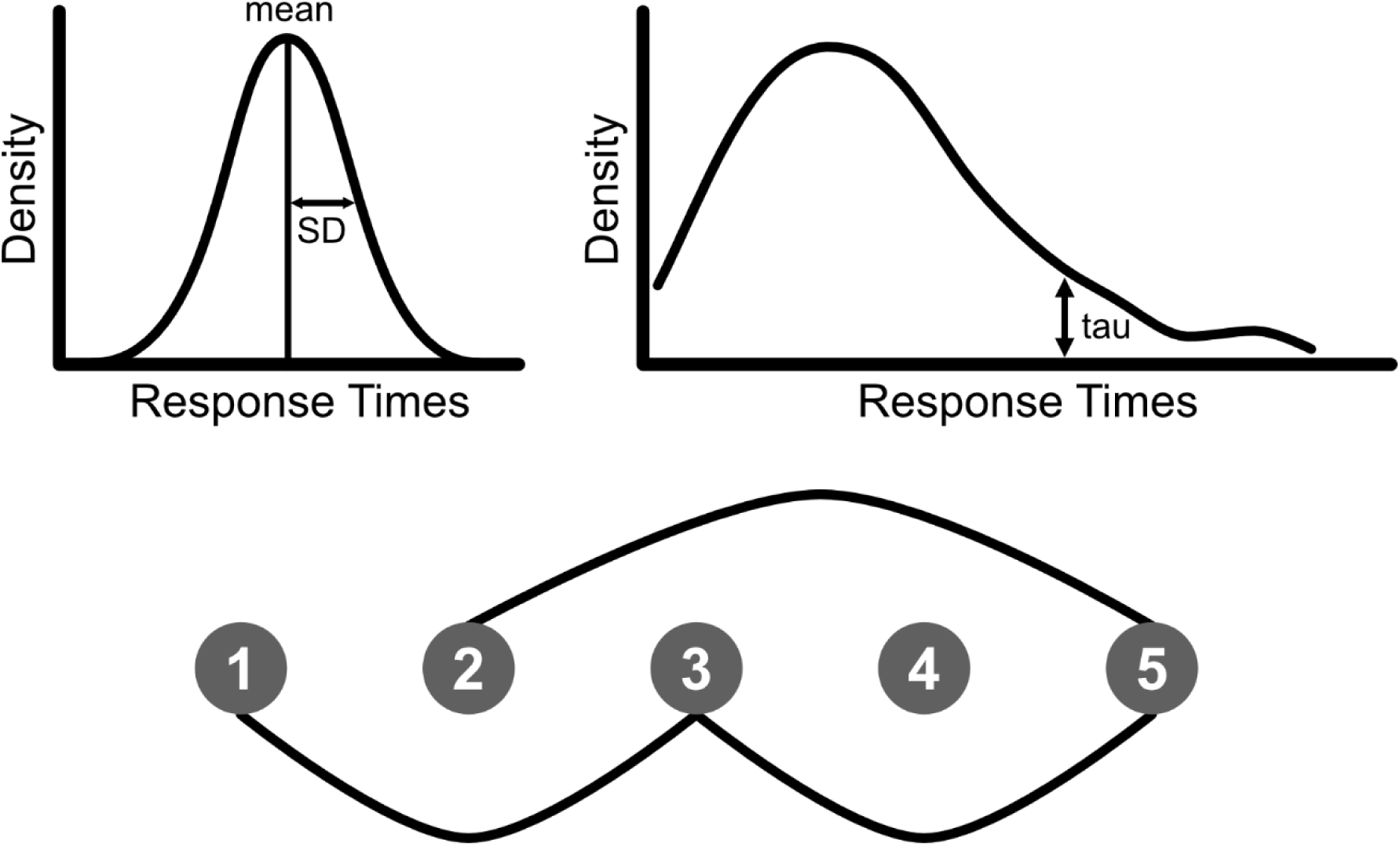
Illustrations of the three intra-individual variability in response time (IIVRT) measures showing (top left) the assumed gaussian RT distribution for standard deviation, (top right) which ex-gaussian RT distribution component is captured by tau, and (bottom) nodes and edges in the graph theory analysis. An *edge* is created (black line) between two *nodes* (black circles 1-5) if they are sufficiently similar to each other within a set threshold and within the same time window (e.g. five nodes in the figure). Each node corresponds to a Go RT.

The graph theory principles and similarity graph algorithm used in the current study were based on the original publication of this method (Fasmer et al., 2018). We applied a nonlinear heuristic algorithm (Fasmer et al., 2018; Kvadsheim et al., 2022) to transform the time series of Go RT data (extracted from the dataset in the order they were completed) into a similarity graph. Every Go RT in the time series was represented by a node in the graph. Using a sliding time window, every RT (index node) was analyzed in relation to a number of neighbour nodes of RTs (Figure 2C); ranges of ± 2 and ± 10 neighbours/trials were applied in the current study. An edge between two nodes indicated that the nodes fulfilled the criterion of similarity (i.e. the difference between RTs was below a predefined threshold). A threshold for similarity of 5% was used in the current study. Greater similarity is reflected by a larger number of edges. Overall, graphs with a higher number of similar Go RTs are described as having higher inter-relatedness on a trial-by-trial basis.

### Statistical Analysis

A mixed-effects, repeated measures analysis of variance (ANOVA) was run on all dependent measures from Go trials. Average RT, SD, tau and number of edges for 2 and 10 neighbours were analysed with a 2 Group (HC, PD) X 2 Side (Left, Right) design. Effect sizes are reported for all significant ANOVA results and statistical significance is set at α = 0.05. Post hoc *t* tests were used to investigate significant ANOVA results. Values are reported as mean ± standard error (SE).

## Results

### Average Response Time

On average, HC and PwPD had comparable Go trial performance. For average RT, there was no effect of Group (F_1,113_ = 0.062, p = 0.804) but a main effect of Side (F_1,113_ = 27.235, p < 0.001, ƞ_p_^2^ = 0.194) with the right index finger RT (821 ± 3 ms) being faster than the left (835 ± 3 ms). Given the predominantly right-handed sample of participants, such speeding of the dominant side is not surprising and has been shown previously in young healthy adults (Coxon et al., 2007). The Group X Side interaction approached significance (F_1,113_ = 3.845, p = 0.052, ƞ_p_^2^ = 0.033) but did not show a trend towards decomposing meaningfully according to group effects, as the difference between groups in left (p = 0.357) and right RT (p = 0.104) did not reach significance.

### Intraindividual Variability – Average Measures

Despite a comparable mean, PwPD showed greater overall variability in their average RT distribution. For standard deviation, there was a main effect of Group (F_1,113_ = 14.337, p < 0.001, ƞ_p_^2^ = 0.113, Figure 3 left) as PwPD exhibited larger SD (64 ± 3 ms) compared to HC (45 ± 4 ms). There was no Group X Side interaction (F_1,113_ = 0.043, p = 0.836) but a trend towards a main effect of Side (F_1,113_ = 3.533, p = 0.063, ƞ_p_^2^ = 0.030) as the left RT showed slightly higher SD (55 ± 3 ms) than the right (53 ± 3 ms).

**Fig. 3.**
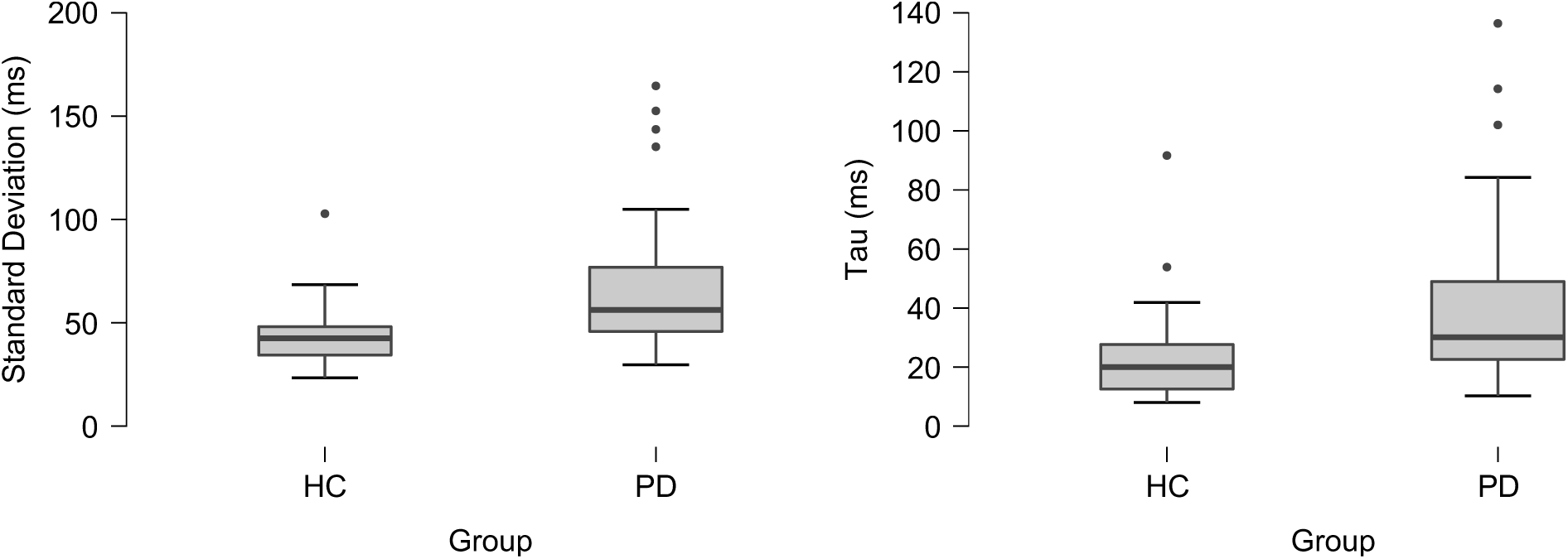
Group effects on intraindividual variability shown via average measures of standard deviation (left) and tau (right). Parkinson’s disease (PD) participants showed greater variability than healthy controls (HC) on average across the entire experimental response time distribution and across slower response times.

In addition to greater overall variability, PwPD also exhibited a greater positive skew of their RT distributions compared to controls. Tau also showed a main effect of Group (F_1,113_ = 8.640, p = 0.004, ƞ_p_^2^ = 0.071, Figure 3 right) as PwPD had larger tau (38 ± 2 ms) than HC (24 ± 4 ms). The main effect of Side reached significance (F_1,113_ = 3.987, p = 0.048, ƞ_p_^2^ = 0.034) with the left RT distribution across groups showing a larger tau (33 ± 3 ms) compared to right (29 ± 2 ms). There was no Group X Side interaction (F_1,113_ = 052, p = 0.819).

### Intraindividual Variability – Trial-by-trial Measures

Across shorter time windows, PwPD showed a reduced degree of trial-by-trial inter-relatedness between RTs. The average number of edges for ± 2 neighbour nodes produced a main effect of Group (F_1,113_ = 11.479, p = 0.001, ƞ_p_^2^ = 0.092, power = 0.98; Figure 4 left) due to PwPD data producing a smaller number of edges (0.59 ± 0.02) compared to HC (0.72 ± 0.03). There was no effect of Side (F_1,113_ = 1.497, p = 0.224) or Group X Side interaction (F_1,113_ = 0.132, p = 0.717).

**Fig. 4.**
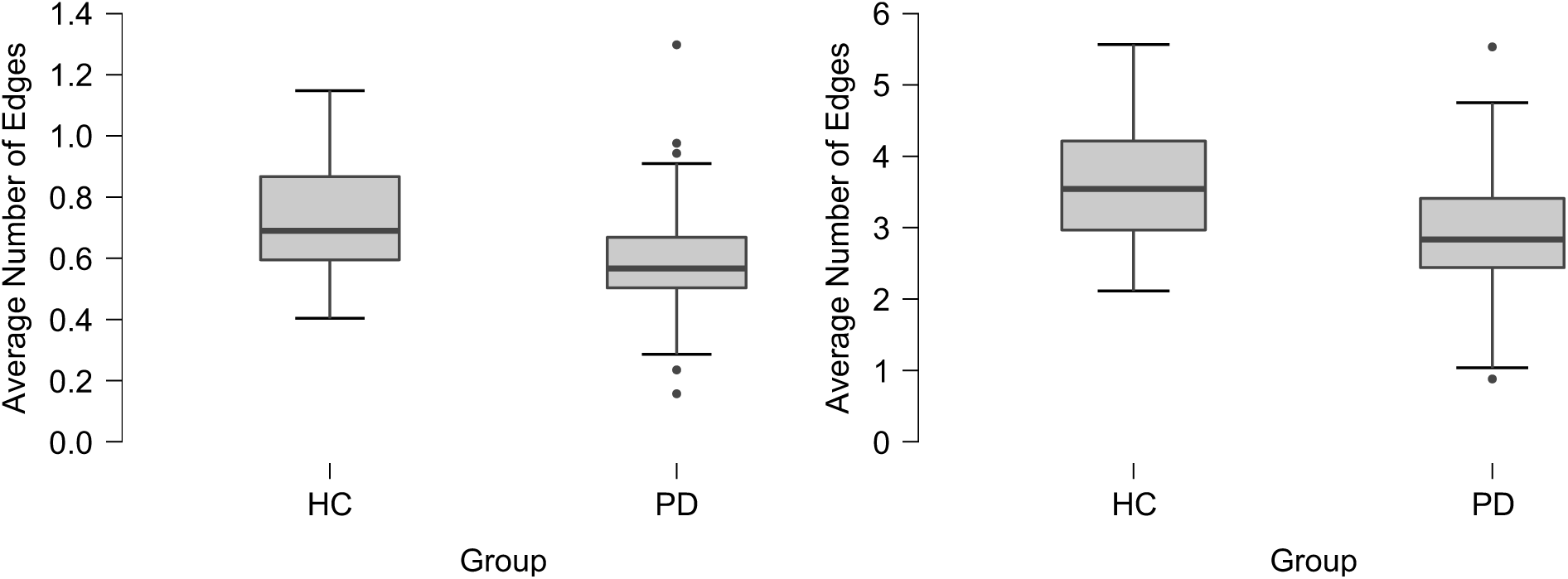
Group effects on intraindividual variability shown via trial-by-trial measures from graph theory analysis. Parkinson’s disease (PD) participants showed reduced response time similarity (smaller number of edges) compared to healthy controls (HC) across time windows of ± 2 trials (left) and ± 10 trials (right).

PwPD also showed a reduced degree of inter-relatedness between RTs across longer time windows. Similar to the ± 2 neighbour analysis, analysing time windows of ± 10 neighbour nodes also produced a main effect of Group (F_1,113_ = 18.794, p < 0.001, ƞ_p_^2^ = 0.143, power = 0.99; Figure 4 right). Data from PwPD showed a reduced number of average edges (2.90 ± 0.09) compared to HC (3.65 ± 0.15). There was no effect of Side (F_1,113_ = 1.034, p = 0.311) or Group X Side interaction (F_1,113_ = 0.099, p = 0.754).

## Discussion

The current study constitutes the first use of the graph theory approach on RT data. As hypothesized, PwPD showed higher trial-by-trial IIVRT on Go trials of the ARIT compared to healthy older adults, indexed via reduced similarity of RTs. These novel results indicate that graph theory may be a useful tool to capture the more rapidly varying changes in RT behaviour that result from the dysfunctional dopamine bursting dynamics present in PD. As predicted, PwPD also exhibited larger IIVRT-tau values compared to controls. This indicates PwPD had more disproportionally longer Go RTs. Consistent with previous findings, we also found PwPD showed greater overall IIVRT-SD compared to controls. These differences in IIVRT between groups cannot be explained simply by worse overall Go trial performance, as average Go RT was comparable between groups. Instead, the IIV findings specifically reflect impaired consistency in performance for PwPD. The reduced trial-by-trial and average RT consistency for PwPD on our behavioural task is therefore likely due to candidate underlying mechanisms such as impaired time perception, motor deficits and/or cognitive impairments. Overall, the current study provides strong evidence for the inclusion of graph theory and ex-gaussian measures of tau, along with the ARIT, in future studies investigating RTs in dopamine-related conditions like PD.

PD was associated with rapidly varying RT behaviour. PwPD had reduced moment-to-moment consistency in their responses compared to healthy controls as captured via the graph theory analysis. Graph theory is therefore a plausible method for addressing the need to isolate particular components of RT variability for more in-depth characterization of IIV (Tamm et al., 2012). Rapid fluctuations in behaviour are likely linked to rapidly changing activity in underlying neural circuitry. As such, the current results suggest that graph theory is a viable tool to capture the high frequency variability in behaviour resulting from dysfunctional dopamine bursting activity in PD. However, it is possible that cerebellar dysfunction (Li et al., 2023) could also be contributing to the inconsistent cognitive-motor performance in the PD group. Cerebellar dysfunction could be interacting with nigrostriatal bursting activity through cortico-cerebellar-thalamo-cortical pathways, specifically via dentate nucleus output to the striatum (Hoshi et al., 2005). The two mechanistic possibilities are not mutually exclusive and warrant further investigation.

People with PD exhibited a greater number of slower responses compared to controls. We are the first study to report higher IIVRT-tau for PwPD relative to controls. Within ADHD literature, increased tau is speculated to reflect failures of sustained attention (Epstein et al., 2011; Leth-Steensen et al., 2000) possibly caused by intrusion of task-negative brain network activity (e.g. default mode network) during task performance (Tamm et al., 2012). Others suggest tau is indicative of intentional cognitive processes (Kieffaber et al., 2006) or should not be interpreted in terms of specific cognitive processes at all (Rieger & Miller, 2020). The interpretation of increased tau for PD is even more uncertain. Our current results are unable to tease apart cognitive deficits from those resulting from difficulties overcoming increased motor inhibition from basal ganglia pathway imbalance in PD (Calabresi et al., 2014). In theory, difficulties with purely motor control could lead to occasionally later RTs and increased tau, although slower motor speed has been unable to explain increased IIVRT in PD previously (de Frias et al., 2007). Overall, despite a currently unknown mechanism, our results show a lack of consistency in responses for PwPD.

People with PD did not experience cognitive-motor problems which disrupted basic performance on our task. This was evident from average Go RT not being significantly different between groups. The comparable performance of the default behavioural response shows patients were performing the task with equivalent processing speed and basic executive function. However, the presence/absence of a difference in average RT between groups is task dependent. Simple reaction time tasks have produced slower (Morrison et al., 2021) and comparable (Camicioli et al., 2008; de Frias et al., 2007) RTs in PD. Similarly, studies using tasks that include a speeded decision making component, such as the Eriksen Flanker (Cagigas et al., 2007; Pappa et al., 2021) and choice reaction time tasks (Camicioli et al., 2008; de Frias et al., 2007; Dujardin et al., 2013), have reported differing average RT results. On the other hand, the increased IIVRT-SD for PwPD seen in our study has been consistently reported across RT tasks (Burton et al., 2006; Camicioli et al., 2008; de Frias et al., 2007; Dujardin et al., 2013; Morrison et al., 2021). Amongst RT tasks the ARIT uniquely requires the correctly timed execution of an internally generated response, rather than an externally cued response. Time perception, especially within the sub-second range (Nani et al., 2019), relies on striatal function (Harrington et al., 2010; Meck et al., 2008) and is therefore impaired in PD (Zhang et al., 2016). Of most relevance for this study, PwPD exhibit comparable averages but higher variability for time estimates compared to healthy controls (Singh et al., 2021). It is therefore possible that impairments in predictive timing processes could be contributing to the larger IIVRT observed for PwPD using the ARIT.

There are two important considerations when interpreting the current results. Firstly, despite the two groups completing the same behavioural task, data for healthy controls and PwPD were collected and compared across two separate studies. It is possible the different experimental environments contributed to the behavioural differences between groups. However, the non-significant group difference for average Go RT is an important factor which argues against this possibility. Nevertheless, a replication of these findings within a single study may be prudent. Secondly, the focus of this study has been on Go trials embedded within an inhibitory control task, as done previously (Epstein et al., 2023). It therefore cannot be ruled out that IIV was influenced by mechanisms engaged during Stop trials that affected behaviour on the subsequent Go trial. However, the vast majority of Go trials were not preceded by a Stop trial so any effect would be minimal. More importantly for the main findings of this study, both groups performed the same combinations of trials.

### Conclusion

PwPD showed comparable average performance but impaired consistency in their Go trial responses on the ARIT compared to controls. Indexing IIV via nonlinear measures from graph theory and tau specifically revealed more rapidly varying trial-by-trial behaviour and a larger proportion of slower RTs, respectively. These characteristics of IIV changes in PD could be reflecting dysfunctional dopamine bursting dynamics, cerebellar dysfunction, cognitive deficits such as attentional lapses, and/or temporal processing difficulties. The current study is the first to use the graph theory approach on RT data and shows it to be a viable method to assess IIV via short-term fluctuations on a trial-by-trial basis. Our study provides strong evidence for the inclusion of graph theory and ex-gaussian measures of tau, along with the ARIT, in future studies to investigate RT behaviour in PD and other dopamine-related conditions.

## Research Transparency Statement

### General Disclosures

#### Conflicts of interest

All authors declare no conflicts of interest.

#### Funding

Open access funding provided by the University of Bergen.

#### Artificial intelligence

No artificial intelligence assisted technologies were used in this research or the creation of this article.

#### Ethics

All data was collected in studies which received approval from local ethics boards (University of Auckland Human Participant Ethics Committee ID: 10083; Health and Disability Ethics Committee ID: 13/NTA/215; University of Birmingham Ethical Review Committee ID: ERN_18-2077AP1A).

### Study Disclosures

#### Preregistration

None.

#### Materials

Code to run the behavioural tasks is publicly available (<u>github repository</u>).

#### Analysis scripts

All analysis scripts are publicly available (<u>github repository</u>, <u>github repository</u>).

## Data Availability

All data produced in the present study are available upon reasonable request to the authors. Materials: Code to run the behavioural tasks is publicly available (https://github.com/hayleymacdonald3/Manuscript_RT-fluctuations-in-PD). Analysis scripts: All analysis scripts are publicly available (https://github.com/hayleymacdonald3/Manuscript_RT-fluctuations-in-PD, https://github.com/erlfas/SimilarityGraph).

https://github.com/erlfas/SimilarityGraph

## Notes

### Competing Interest Statement

The authors have declared no competing interest.

### Author Declarations

All data was collected in studies which received approval from local ethics boards: University of Auckland Human Participant Ethics Committee ID: 10083; Health and Disability Ethics Committee ID: 13/NTA/215; University of Birmingham Ethical Review Committee ID: ERN_18-2077AP1A.

